# Adrenergic nerve fibers are depleted in adenomyosis myometrium

**DOI:** 10.64898/2026.11.16.26368015

**Authors:** Nicolas R. Chevalier, Emilie Huon, Olympe Ségard, Mariam Ouali, Marine Plancke, Clara Boer Wigman, Thierry Savy, Thomas Pujol, Pierre Bourdoncle, Gauthier Dietrich, Erick Petit, Ylann Abrahami, Jordan Kouby, Julien Adam, Marie-Charlotte Lamau, Carole Abo

**Affiliations:** Laboratoire Matières et Systèmes Complexes, Université Paris Cité, CNRS, F-75013 Paris, France; Institut Cochin, INSERM U1016, CNRS UMR 8104, Université de Paris (UMR-S1016), Paris, France; Service de Chirurgie Gynécologique, Hôpital Paris Saint-Joseph, 75014 Paris, France; Service d’Anatomo-cyto-pathologie, Hôpital Paris Saint-Joseph, 75014 Paris, France

**Author notes:** **Corresponding author:** Dr. Nicolas R. Chevalier, CNRS researcher, Intestinal & Uterine physiology, Laboratoire MSC, 10 rue Alice Domon, 75013 Paris, France. **Funding Statement** : This study was funded by the Fondation pour la Recherche sur l’Endométriose, project HYPERU, under the auspices of the Fondation pour la Recherche Médicale, project FRE202212016469. **Disclosure Statement:** All authors declare that they have no competing interests. **Attestation Statement:** Data regarding any of the subjects in the study has not been previously published unless specified. Data will be made available to the editors of the journal for review or query upon request. **Data Sharing Statement:** Essential data generated or analyzed during this study are included in this report and its supplementary information files. Further data are available from the corresponding author upon reasonable request. **Trial registration:** Approval for this study was granted on the 27^th^ of January 2025 under IRB number 00012157 from the Groupe Ethique Recherche Médicale (GERM) of Hôpital Paris Saint Joseph. The first patient was included on the 26^th^ of February 2025. **Capsule:** The myometrium of adenomyosis patients present a significantly lower amount of muscle-relaxing, adrenergic nerve fibers. We explain how this underpins junctional zone thickening and dyscontractility in this pathology.

**Keywords:** histology, sympathetic innervation, uterine contractility, junctional zone, second harmonic generation microscopy

## Abstract

**Objective:** Uterine dyscontractility in adenomyosis and endometriosis is at the heart of pain symptoms and disease pathogenesis. To elucidate possible causes of dyscontractility, we examined the distribution of smooth muscle, blood vessel, collagen and nerve fibers in full-thickness, serosa-to-endometrium myometrium of adenomyosis patients versus control.

**Design:** Retrospective study.

**Subjects:** Uterine biopsies from patients who underwent hysterectomy for adenomyosis (n = 26) or other non-adenomyosis indications (n=22, mostly myomas).

**Main Outcome Measures:** Density profiles along the serosa-to-endometrium axis of smooth muscle (αSMA), endothelium (PECAM), general innervation (PGP9.5) and adrenergic innervation (TH) obtained by immunofluorescence, mosaic epifluorescence imaging and machine-learning assisted segmentation. Fibrous collagen type I&III was quantified by second-harmonic generation microscopy (SHG).

**Results:** The density profiles of smooth muscle, endothelium, collagen and PGP9.5 were similar between both patient groups. Sympathetic adrenergic nerve fiber density radiated from the serosa inwards, with a continuously decreasing density from the outer to the inner myometrium. Adrenergic inhibitory nerve fibers were 4 times less dense in the inner myometrium of adenomyosis uteri compared to controls.

**Conclusion:** The enrichment of inhibitory adrenergic nerve fibers in the outer vs inner myometrium provides a neuro-muscular mechanism by which contractions are restricted to the inner myometrium outside of pregnancy. The depletion of TH fibers in adenomyosis implies that a higher thickness of myometrium is involved in uterine contractions, thus explaining junctional zone thickening and dyscontractility in this pathology. This depletion may be due to defective post-pregnancy adrenergic re-innervation of the myometrium.

## Introduction

The myometrium is the thickest smooth muscle (SM) of the human body: ∼2.5 cm in the non-pregnant female. Its menstrual-cycle-dependent peristaltic contractions (De Ziegler et al., 2001) play a major role in sperm propulsion at ovulation (Kunz et al., 1996) and in menses evacuation (Lyons et al., 1991). While contraction of the full thickness of the myometrium is required for parturition, only the inner myometrium participates in peristaltic activity of the non-pregnant uterus (1–3). This implies that the outer myometrium is actively maintained in a quiescent state outside of pregnancy. In endometriosis patients, intra-uterine pressure, contraction frequency and amplitude are elevated by a factor 2-3 compared to healthy women (Bulletti et al., 2002). This hypercontractility likely plays a causal role in disease emergence, by favoring chronic retrograde transport of menstrual debris to the abdominal cavity (Kunz et al., 2005; Sampson, 1927; Vercellini et al., 2024). Adenomyosis is characterized by the invasion of endometrium inside the myometrium (4). Poor myometrial contraction coordination has recently been reported in women suffering from this pathology (Rees et al., 2024). A thickened junctional zone (JZ >12 mm) on MRI is 85-93% specific of adenomyosis (5,6). JZ thickening is also found in 79% of endometriosis patients (7).

The reasons underlying JZ thickening and dyscontractility in endometriosis and adenomyosis remain unknown. One study (8) found increased α-SMA staining and collagen type-I labeling in the inner myometrium of adenomyotic vs non-adenomyotic uteri, but this was not confirmed in two other independent studies (9,10). Doppler sonography has suggested potential alterations to myometrium vasculature in adenomyosis (6), with conflicting reports of hyper-(11) or hypo-vascularization (12). One investigator reported the absence of PGP9.5 nerve fibers around adenomyosis lesions (13). Other investigations of innervation in adenomyosis have focused mostly on the endometrium (14–16).

In spite of the organ-level phenomenology of the JZ, histological studies rarely examine the entire thickness of the uterine wall (9,17,18), and our general knowledge of the myometrium anatomy remains lacunar. We therefore use here modern technics of mosaic epifluorescence imaging combined with machine-learning assisted segmentation to quantitatively characterize the cellular organization of the full-thickness myometrium and its alterations in adenomyosis. We present serosa-to-endometrium density profiles of smooth muscle, vasculature, general innervation (PGP9.5), and sympathetic adrenergic innervation (TH) in control and adenomyosis patients. We also harness label-free second harmonic generation imaging (SHG) (19,20) to quantify collagen I & III fibers; this powerful tool has only recently been introduced in the field of gynecology (21). We uncover a sharp depletion of adrenergic muscle-relaxing nerve fibers in the inner myometrium of adenomyosis patients compared to control patients. This finding rationalizes uterine dyscontractility and JZ thickening in adenomyosis.

## Materials and Methods

### Study design and patient cohort

48 human myometrium strips were obtained from women following hysterectomy at Hôpital Paris Saint Joseph (Table S1). Approval for this study was granted under IRB number 00012157 from the Groupe Ethique Recherche Médicale (GERM) of Hôpital Paris Saint Joseph. All women were provided with written information about the study and consented to participate.

### Tissue collection and preparation

After surgery, the uterus was opened by sectioning the anterior wall along the cervix-to-fundus axis. A ∼25×10×10 mm block of myometrium, from serosa to endometrium, was cut out from the anterior uterine wall, at a position halfway between cervix and fundus, avoiding, when present, any visible myomas. The tissue block was immediately fixed in 4% PFA overnight, at 4°C. It was then washed 3×1 hour in PBS, and kept in PBS with 0.02% sodium azide at 4°C until sections were performed. The tissue block was dehydrated in 30% sucrose solution for 24h and embedded in PolyFreeze (Sigma). Biopsies were oriented in a way that the presumably serosal longitudinal SM fibers (22,23) were in the section plane, and that presumably circular fibers were perpendicular to the section plane. Some samples where orientation was lost or unknown were not included in the SM fiber orientation analysis. 18 µm-thick sections were cut on a cryotome (Leica) and adhered on SuperFrost glass slides.

### Immunofluorescence and imaging

Permeabilization and blocking were performed in PBS + 1% bovine serum albumin (Fraction V, Sigma) + triton 0.1% for 1 hour. Slides were incubated with the following antibodies overnight: αSMA Cy3-conjugated (Sigma C6198, 1:1000), PECAM AF647-conjugated (SCBT sc-376764, 1:100), rabbit anti-PGP9.5 (Abcam AB108986, 1:100) or rabbit anti-TH (Abcam AB112, 1:100). When a secondary antibody was necessary (PGP9.5, TH), we washed the slide 3×10 min in PBS and then applied anti-rabbit Cy3 secondary (Thermofisher, 1:500) for 2 h. All steps were performed at RT, under constant, gentle agitation to optimize staining homogeneity. All slides were finally washed in PBS, coverslipped, and imaged.

### Microscopy

Imaging was performed at the ImagoSeine microscopy platform on an inverted Zeiss wide-field epifluorescence microscope, at x10 magnification, by mosaic-stitching of ∼100-300 tiles, depending on sample size. SHG is a multiphoton, label-less method to image collagen I&III (24). It was performed with an upright Leica SP8 DIVE FALCON (Wetzlar, Germany) coupled with a pulsed laser controlled by LAX software on IMAG’IC facility. SHG (excitation: 800 nm, emission: 389-412nm), two-photon autofluorescence (ex: 800 nm, em: 501-553 nm), at x10 in mosaic mode. Acquisition settings were kept constant for all samples in order for collagen densities to be quantitatively compared.

### Image analysis

Segmentation was performed using the machine-learning open-source program Ilastik version 1.4.2rc1 (25). For each staining, a new model was trained until the segmented image visually matched the distribution of the labeled cells. The segmented binary was exported and its profile measured along a curvilinear line, 1 to 5 mm thick, from serosa to endometrium. The result was converted in % area density, all profiles were subsampled to 1000 points ranging from 0 (serosa) to 1 (SM-endometrium junction) and average curves for each patient group produced under Matlab.

## Results

The characteristics of the patients included in this study are shown in Table S1. n=26 patients presented with adenomyosis as confirmed by MRI exams. n=22 patients presented mostly with myomas (80%) or other non-adenomyosis indications (20%). All biopsies were collected from the anterior wall of the uterus, at a position halfway between cervix and fundus. We aimed for n≥20 density profiles for each marker (5) and for each patient group (2), retrieving a total of 213 profiles (Table S2). All samples were subject to the same experimental and machine-learning assisted segmentation pipeline (Fig. S1). For each marker, we show representative post-segmentation images across the full myometrium thickness, magnified images in the outer (OM) and inner (IM) myometrium, and average density profiles of control and adenomyosis groups. Individual patient profiles are provided as supplementary material (Fig. S2-S6).

Smooth muscle area density increased from ∼50% in the OM to ∼60% in the IM (Fig.1A, Fig.2A,D,G), consistent with previously reported SM-to-connective-tissue ratios (18). SM density profiles of control and adenomyosis patients were almost identical (Fig.2G). Distinct orientations of SM fibers were visible in this order from serosa to endometrium: (I) an outer serosal layer, (II) an inner serosal layer whose fibers were always perpendicular to the outer serosal layer, (III) a thick middle layer with intertwining, probably spirally arranged fibers (22,23), and (IV) an inner layer adjacent to the myometrium. Surprisingly, we found considerable variation from one uterus to the other in the absolute orientation of the outermost (I) and innermost (IV) layers (Table S3). SM orientation differences were however not correlated with pathology.

**Figure 1.**
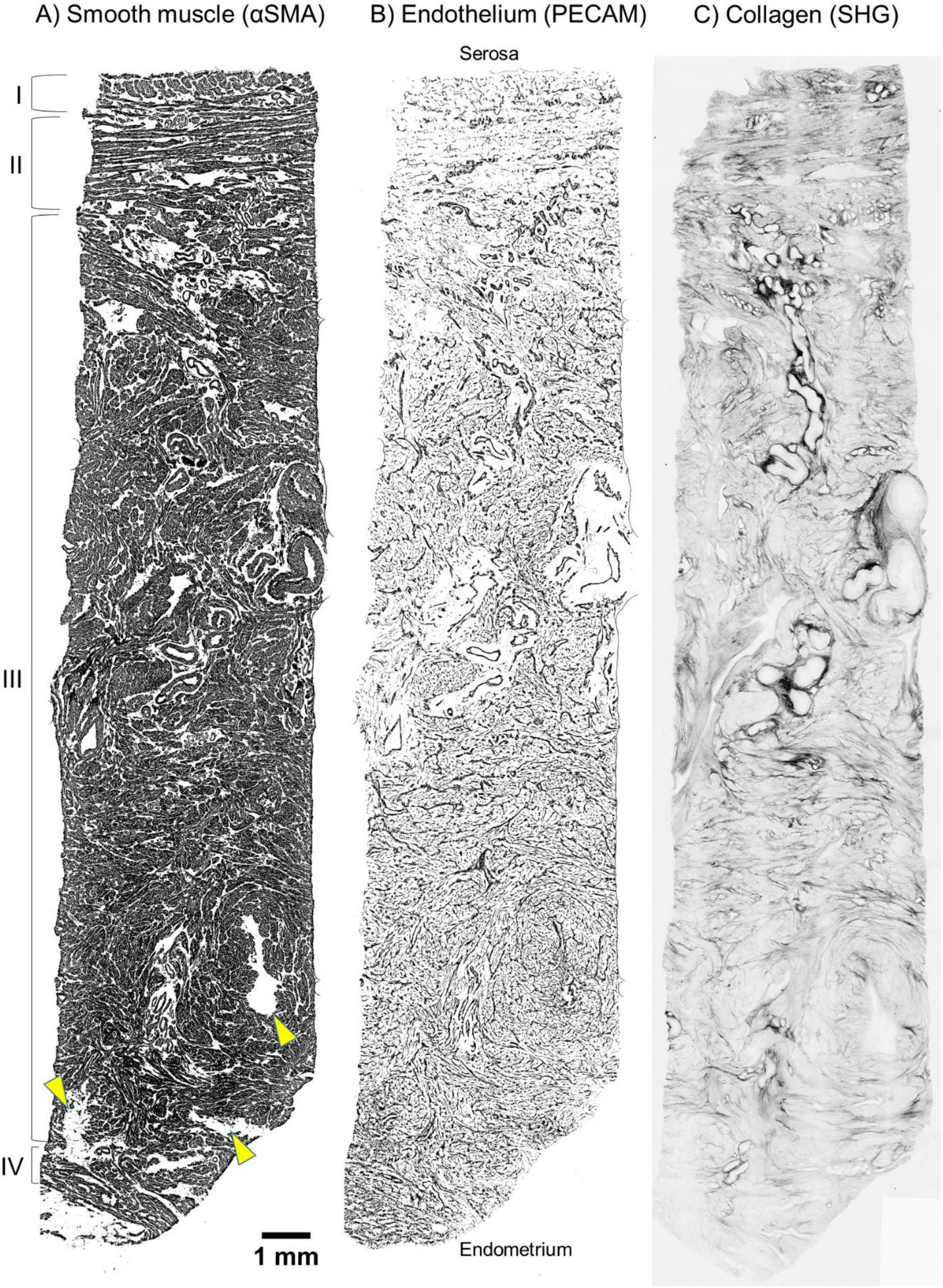
Smooth muscle, blood vessel and collagen distribution in full-thickness myometrium sections. These representative distributions are from an adenomyosis patient. The serosa is at the top, the endometrium at the bottom. Yellow arrowheads point at three groups of adenomyosis lesions. A) Post-segmentation α-SMA binary: the muscle density increases from serosa to endometrium, and layers with distinct orientations (I-IV) are visible. In this example fibers are circular (I), longitudinal (II), spiral (III) and longitudinal (IV). B) Post-segmentation PECAM binary: the capillary density increases from serosa to endometrium. C) Native SHG signal: collagen content decreases from serosa to endometrium. The scale bar is identical for the three images.

**Figure 2.**
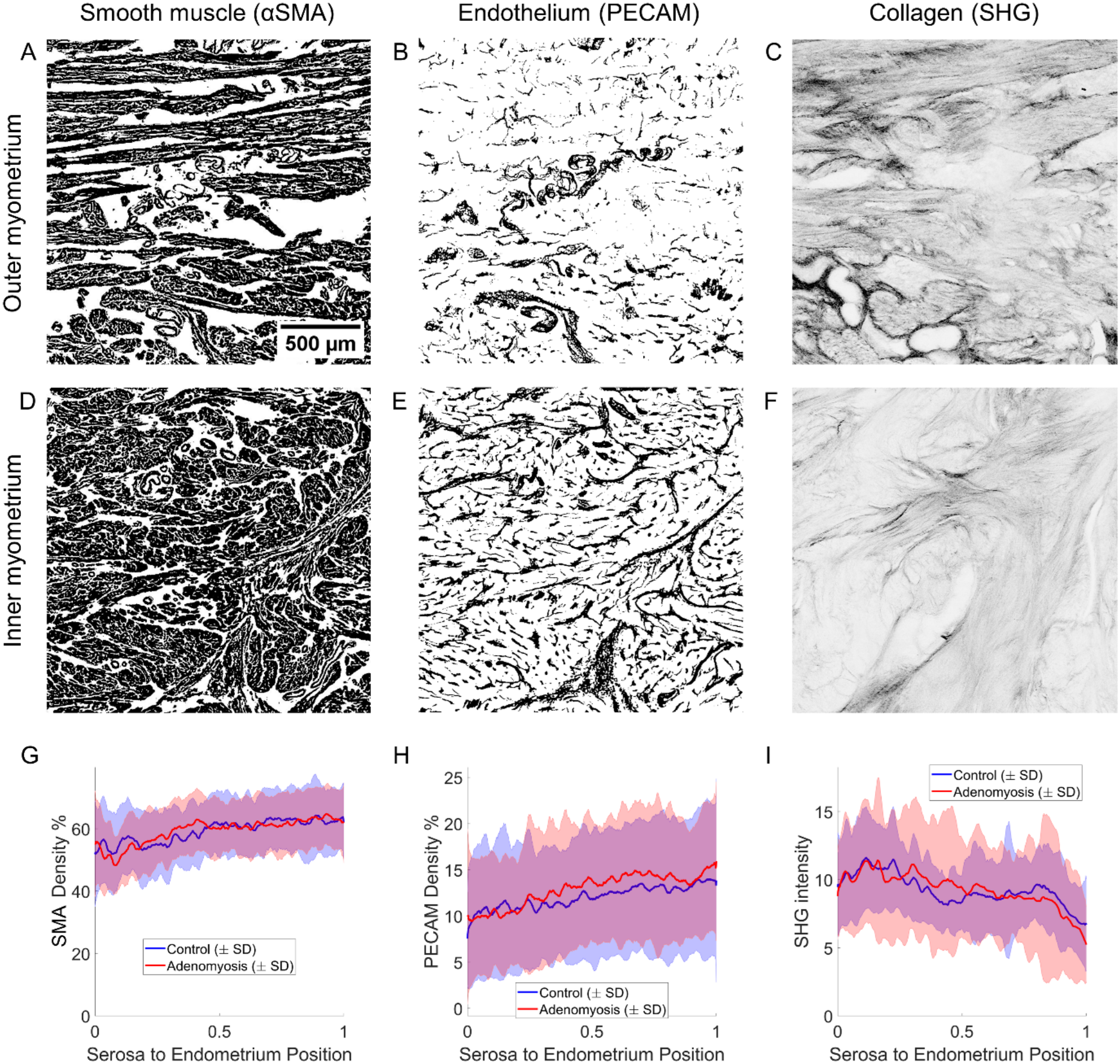
Smooth muscle, blood vessel and collagen density profiles are similar in control and adenomyosis patients. (A-F) Magnified regions of Fig.1. The scale bar is the same for all images. (G) SMA density profile, n=20 controls, n=23 adenomyosis. (H) PECAM density profile, n=20 controls, n=24 adenomyosis. (J) SHG intensity profile, n=20 controls, n=20 adenomyosis. SHG intensity is proportional to fibrous collagen content. The thicknesses of the different myometria were normalized, with the position of the serosa=0 and the SM-endometrium junction = 1.

Vasculature, as quantified by the density of endothelial PECAM positive cells, increased from ∼10 % in the OM to ∼15% in the IM. Since PECAM only labels the endothelial lining, it does not directly yield the blood vessel area density, which should additionally include blood vessel lumen and tunica (Fig.S7). Capillary vessels were most dense in the IM, medium sized vessels in the OM, while very large vessels were usually found towards the middle of the myometrium (Fig.1B, Fig.2B,E). PECAM density profiles from control and adenomyosis patients were similar (Fig.2H).

Collagen was imaged by SHG microscopy, together with GFP autofluorescence. SHG directly outputs a signal proportional to the amount of fibrous collagen (type I and III). Collagen fibers were present in all interstitial spaces between muscle fibers, and were particularly enriched around large blood vessels. Collagen fiber density decreased by a factor ∼2 from the OM to the IM in both patient groups (Fig.1C, Fig.2C,F,I). Dense collagen fibers, that partially co-localized with strongly autofluorescent fibers (ex: 800 nm, em: 501-553 nm), were particularly present in the OM (Fig.S8). Autofluorescence was likely due to elastin (9,26). Because of this strong and non-homogeneous background GFP autofluorescence, we avoided using any GFP-tagged secondary antibodies. There were no significant differences in the collagen profiles of the two patient groups (Fig.2I).

PGP 9.5 and TH positive fibers were 1-3 µm thick (Fig.3, Fig.S1). We did not find any nerve cell bodies in the myometrium. PGP9.5+ fiber density continuously decreased from ∼1% in the OM to ∼0.5% in the IM (Fig.3A, Fig.4A,D,G). It was lower in adenomyosis patients, but the difference was not statistically significant (p=0.14, Fig.4G). TH density profiles in control patients decreased continuously from ∼0.5% in the OM to ∼0.25% in the IM (Fig.3B, Fig.4B,E,H): adrenergic nerve fibers represented ∼half of all nerve fibers in the myometrium. TH fiber density was lower in adenomyosis than in controls by a factor 1.5 in the OM (p=0.014), and by a factor 4 in the IM (p<0.0001, Fig.3C, Fig.4C,F,I). This depletion was observed even in the myometrium from adenomyosis patients who did not present with lesions in the particular anterior myometrium biopsy that we had collected (n=12/24).

**Figure 3.**
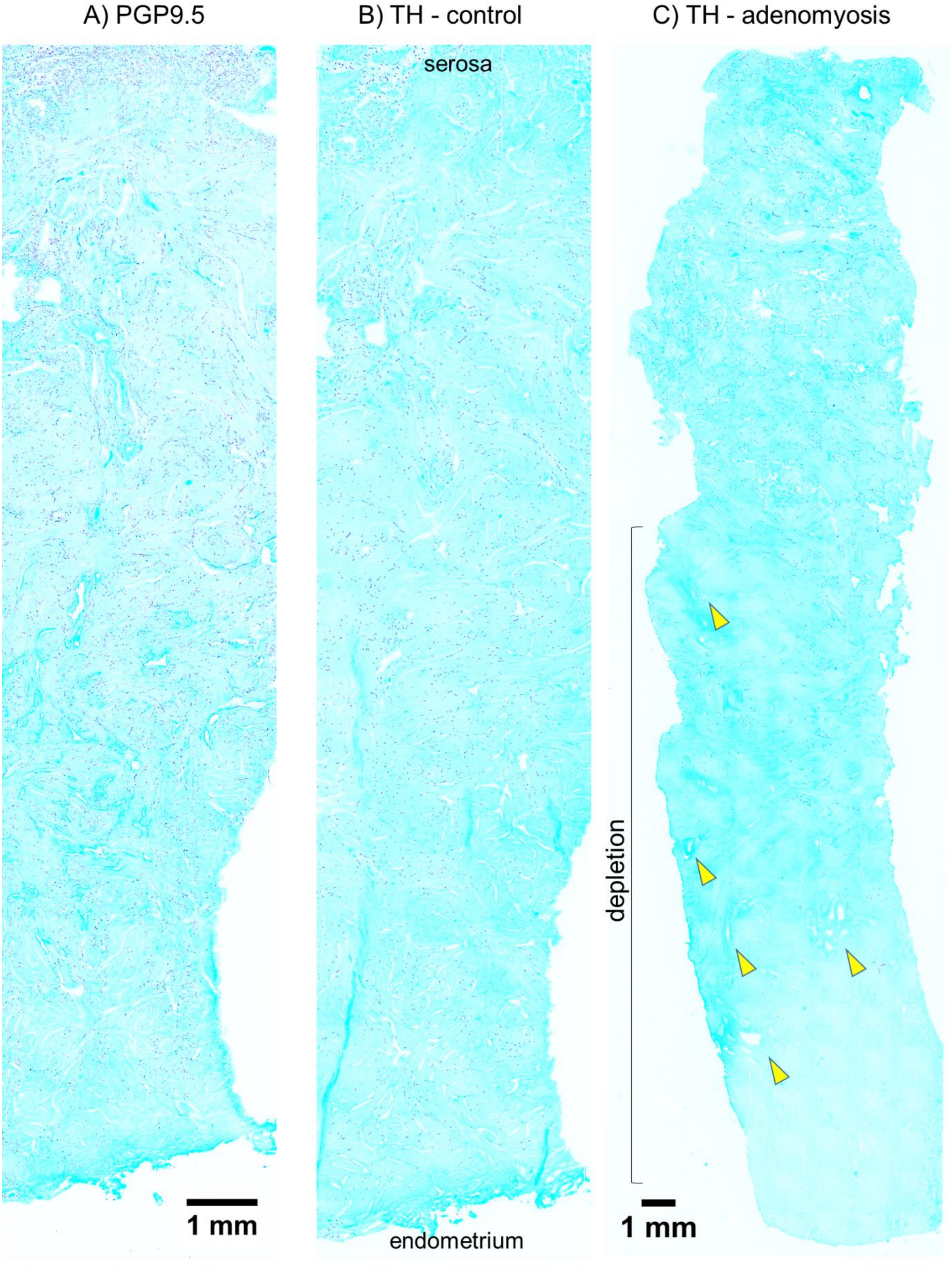
Nerve fiber distribution in full-thickness myometrium sections. Post-segmentation nerve fibers are in dark blue and magenta. The native, low-contrast fluorescence signal is shown in cyan to outline tissue contours. (A) Representative PGP9.5 distribution, here from a control patient presenting with myoma. (B) Representative TH fiber distribution in the same control patient as in (A). (C) Representative TH fiber distribution in an adenomyosis patient, showing depletion in the inner myometrium. Arrowheads point at adenomyosis lesions.

**Figure 4.**
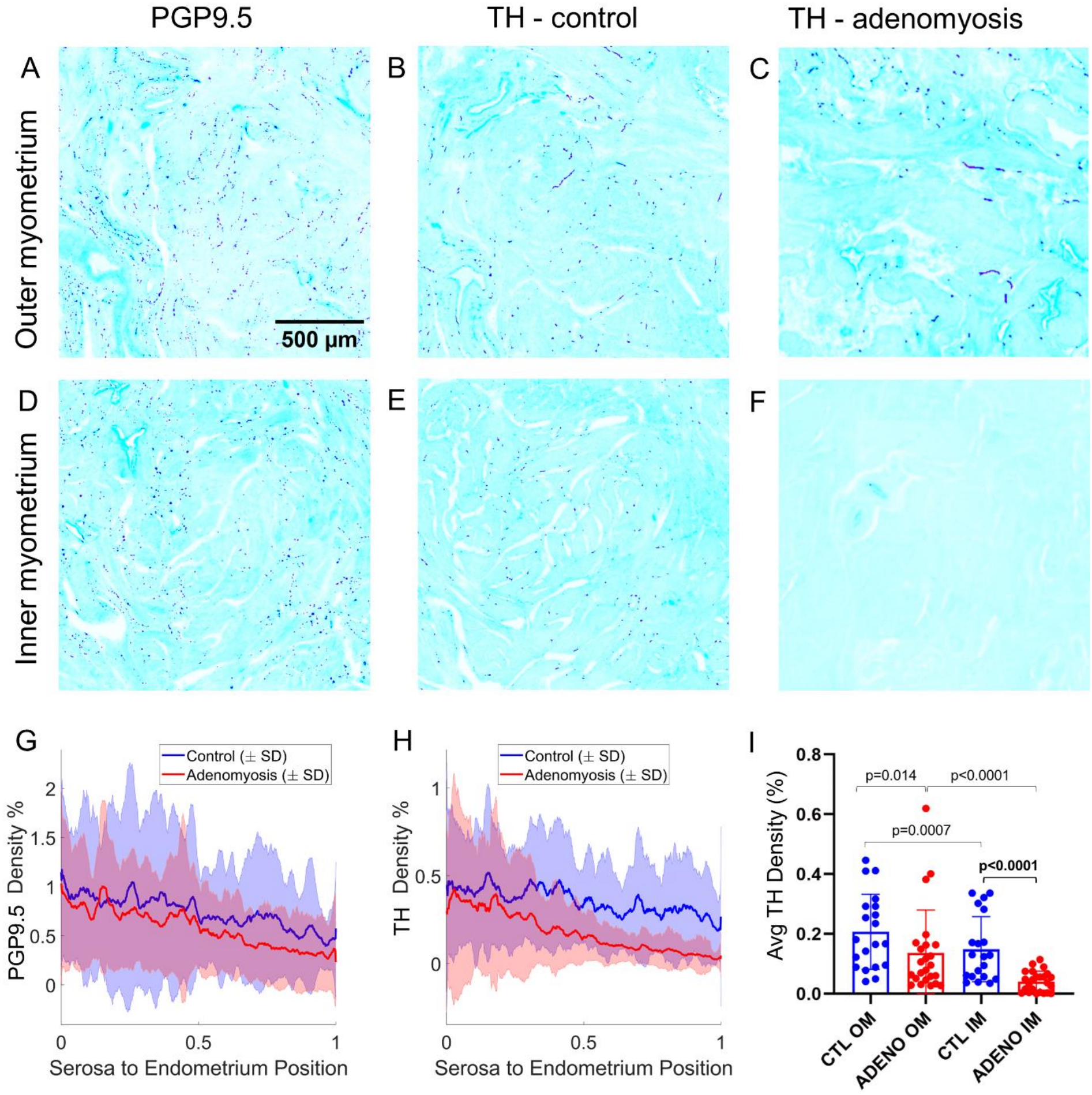
Adrenergic nerve fiber depletion in the inner myometrium of adenomyosis patients. (A-F) Magnified regions of Fig.3. (G) PGP9.5 density profile, n=20 controls and n=22 adenomyosis. (H) TH density profile, n=20 controls and n=24 adenomyosis. (I) Average TH density in the outer myometrium (OM, from 0 to 0.5) and inner myometrium (IM, from 0.5 to 1) for control and adenomyosis patients. Comparisons of the OM and IM were performed using the paired Wilcoxon test; comparisons between control and adenomyosis patients with the unpaired Mann-Whitney test.

## Discussion

### Vascularization, smooth muscle and fiber orientation across the myometrium

The increasing density of SM and capillary vessels from serosa to endometrium is consistent with previous investigations (10,18). SM and blood vessel density profiles were similar between the two patient groups. SM fibers at the serosa are classically described as being longitudinally oriented (22); we found that the outer layer is actually comprised of two sublayers with orthogonal orientations, and that their absolute orientation varied from one patient to the other. In addition, we could only confirm a circular orientation of the innermost layer in about ∼75% of patients. It is possible that SM fiber orientation in the myometrium is affected by stretch (27), which reaches extreme levels during pregnancy. Of note, we found that the innermost layer was ∼2-3mm thick (Table S3); the JZ is about 5-8 mm thick in healthy, menstruating women, and therefore encompasses both the inner layer, and a fraction of the middle spiral layer.

### The outer myometrium has a higher collagen and elastin fiber content

The decrease in collagen content from serosa to endometrium we demonstrate here by quantitative SHG microscopy is consistent with a previous investigation (18). Collagen content is intimately related to tissue stiffness and a higher stiffness of the OM compared to the IM is seen on ultrasound elastography images (28,29). We did not find a difference in the collagen profiles of control and adenomyosis patients, unlike what was found in another study (9). These investigators relied on the measurement of the maximum diameter of fibrils after immunohistochemical staining for collagen I and III; we believe SHG provides a more reliable, quantitative and unbiased measure. Strong GFP autofluorescence at the outer myometrium was likely due to elastin (9,26): we caution other investigators on the use of GFP secondary antibodies in investigating the myometrium.

### The centripetal organization of adrenergic nerves in the myometrium determines the effective thickness of tonic muscle in the non-pregnant uterus

Nerve fibrils from the inferior hypogastric plexus (30) enter the serosa and radiate in fetal human uteri “from outside in” (31). We found the same centripetal organization of nerve fibers in the adult myometrium. We did not observe plexi, i.e. concentration of nerves in particular anatomic planes, as has been reported in the isthmic portion of the uterus (32).

We have recently shown that the myometrium has a uniform organ-bath contractility (33) : IM and OM have similar *ex vivo* contraction amplitude and frequency. Because nerve connections have been severed during the hysterectomy operation, these ex vivo experiments reveal a uniform, spontaneous *myogenic* contractility. When contractility is observed in-vivo by TVUS or MRI, only the IM is seen to contract (1–3): contractile waves fade as they enter the OM. A simple synthetic explanation of these results is that activity of the myometrium results from: 1°) a uniform, spontaneous *myogenic contractility*, 2°) a *nerve-mediated adrenergic inhibition* of this myogenic contractility in the OM. This combined neuro-muscular activity determines *in-fine* the thickness of myometrium that is involved in contractile activity of the non-pregnant uterus. The thickness of tonic SM is precisely the JZ (34,35).

### Functional consequences of adrenergic nerve fiber depletion in adenomyosis

We found similar PGP9.5 density profiles in control and adenomyosis patients. A previous study focusing on the SM-endometrium junction (14) had reached similar conclusions: PGP9.5 innervation density was not correlated to adenomyosis or fibroids, but to the intensity of pain. In contrast, Quinn reported the complete absence of PGP9.5 positive fibers in and around adenomyosis lesions (32); our data indicates slightly lower levels of PGP9.5 in the IM of adenomyosis patients compared to myoma controls.

TH adrenergic fibers were significantly depleted in the IM of adenomyosis uteri. This depletion entails that a greater thickness of muscle is involved in the uterine contractile activity of these patients. This has several functional consequences: 1°) a thicker JZ (34,35), which is today among the diagnostic criteria of adenomyosis, 2°) higher intra-uterine pressure (36), which is closely related to pain sensation (37,38), 3°) the involvement of deeper layers of the myometrium, and in particular the spiral and longitudinal layers which can generate buckling-type deformation of the epithelial lining (39). These deformations are likely initiators of adenomyosis pathogenesis.

### Why are adrenergic fibers depleted in the myometrium of adenomyosis patients?

TH fibers were depleted (<0.1% density) in 21/24 adenomyosis patients, but only 50% of them presented with adenomyosis lesions in the tissue we collected. Moreover, the lesions, when present, generally occupied a minor fraction of the total myometrium area examined. This suggests that TH fiber depletion is systemic: it may be a cause rather than a consequence of the local invasion of the myometrium by uterine epithelium.

Uterine adrenergic innervation is intrinsically plastic (40). Adrenergic nerves degrade during pregnancy, and are absent at parturition (41,42). This is consistent with the idea that only a fraction of the myometrium is required for uterine peristalsis outside of pregnancy, but that its full myogenic potential is necessary to give birth. It also means that adrenergic innervation has to grow back after parturition. Adenomyosis is frequently diagnosed after one or several pregnancies (43). It is therefore possible that adenomyosis could result from defective adrenergic re-innervation post-parturition, with possible links to contraction-induction medication or caesarian section trauma (44–46).

Previous studies have convincingly demonstrated that high estrogen levels reduce the extent of adrenergic innervation (47–49). Contraceptive pills are the number one line of treatment in endometriosis and adenomyosis. Because they reduce the serum concentrations of estrogen by a factor ∼10 (50), they should, theoretically, favor adrenergic innervation and consequently reduce uterine hyper-contractility. Dedicated studies on the effect of the combined pill, of progesterone-only pill, and of pill withdrawal on the adrenergic innervation of the myometrium in animal models would be very beneficial. The plasticity of sympathetic uterine innervation may offer decisive cues to re-establish normal uterine innervation and contractile activity in patients on the long-term.

### Study limitations

We did not have access to patient MRI data, and were thus not able to directly correlate JZ thickness with the extent of TH fiber depletion. Our argument instead relies on the fact that the JZ is statistically thicker in adenomyosis patients compared to controls. We also in most cases (34/48) did not know the menstrual cycle phase of the patient at the time of hysterectomy; we however believe that cyclic myometrium architecture changes should be small compared to the vast TH fiber depletion observed in the adenomyosis group. Finally, we have investigated here only the average density profiles of various cellular components across the myometrium thickness: local remodeling around adenomyosis lesions will be reported elsewhere.

## Conclusions

We examined the density gradients of smooth muscle, blood vessel, collagen and nerve fibers in full thickness myometrium sections. Our study revealed a drastic depletion of inhibitory, adrenergic nerve fibers in the inner myometrium of adenomyosis patients. This depletion entails that a thicker layer of uterine muscle is tonic in these women, explaining their characteristically thick JZ. We hypothesize that the nerve depletion could be due to defective re-innervation of the myometrium post-parturition and/or hyper-estrogenism. Future investigations may reveal how the intrinsic plasticity of adrenergic innervation could be harnessed to re-establish normal uterine contractile activity in adenomyosis, endometriosis and severe dysmenorrhea.

## Data Availability

Essential data generated or analyzed during this study are included in this report and its supplementary information files. Further data are available from the corresponding author upon reasonable request.

## Acknowledgments

We gratefully acknowledge funding by the Fondation pour la Recherche sur l’Endométriose, project HYPERU, under the auspices of the Fondation pour la Recherche Médicale, project FRE202212016469. Emilie Huon was financed by an Ecole Doctorale de Physique de l’Ile-de-France (EDPIF) doctoral fellowship. We thank the anatomopathology staff of Hôpital Paris Saint Joseph for assistance in biopsy retrieval. We thank Thomas Rios, Xavier Baudin and Guillaume Quibeuf from the ImagoSeine imaging platform for help with imaging.

