## Supplementary Material for "Adrenergic nerve fibers are depleted in adenomyosis myometrium"

| | Control group | Adenomyosis group | Unpaired Welch test |
|---|---|---|---|
| **Number of patients** | 22 | 26 | |
| **Age (mean ± SD)** | 46.6 ± 6 | 43.5 ± 6 | p=0.07 |
| **Parity (mean ± SD)** | 1.5 ± 1.0 | 1.6 ± 1.0 | p=0.84 |
| **Hormonal treatment** | 4 [18%] | 7 [27%] | |
| **Menopause** | 3 [14%] | 0 | |
| **Pathology and co-occurrences** | | | |
| ***Myoma*** | 18 [82%] | 4 [15%] | |
| ***Adenomyosis*** | 0 | 30 [100%] | |
| ***Endometriosis*** | 0 | 13 [50%] | |
| ***Adenoma*** | 1 [4%] | 0 | |
| ***Hyperplasia*** | 1 [4%] | 0 | |
| ***Bifid uterus*** | 1 [4%] | 0 | |
| ***Ovarian cancer*** | 1 [4%] | 0 | |
| **Adenomyosis lesions visible on anterior myometrium biopsy** | 0 [0%] | 12 [46%] | |
| **Thickness Anterior Myometrium (mm)** | 23.5 ± 7 | 23.7 ± 6 | p=0.9 |

***Table S1. Characteristics of the two patient groups considered in this study.*** *The majority of control patients underwent hysterectomy for myoma, and none of them had adenomyosis. Half of the adenomyosis patients also presented with endometriosis, and 15% had co-occuring myoma.*

| CONTROL | | | | | | ADENOMYOSIS | | | | | |
|---|---|---|---|---|---|---|---|---|---|---|---|
| Patient | SMA | PECAM | PGP | TH | SHG | Patient | SMA | PECAM | PGP | TH | SHG |
| 1 | | | | | | 1 | | | | | |
| 2 | | | | | | 2 | | | | | |
| 3 | | | | | | 3 | | | | | |
| 4 | | | | | | 4 | | | | | |
| 5 | | | | | | 5 | | | | | |
| 6 | | | | | | 6 | | | | | |
| 7 | | | | | | 7 | | | | | |
| 8 | | | | | | 8 | | | | | |
| 9 | | | | | | 9 | | | | | |
| 10 | | | | | | 10 | | | | | |
| 11 | | | | | | 11 | | | | | |
| 12 | | | | | | 12 | | | | | |
| 13 | | | | | | 13 | | | | | |
| 14 | | | | | | 14 | | | | | |
| 15 | | | | | | 15 | | | | | |
| 16 | | | | | | 16 | | | | | |
| 17 | | | | | | 17 | | | | | |
| 18 | | | | | | 18 | | | | | |
| 19 | | | | | | 19 | | | | | |
| 20 | | | | | | 20 | | | | | |
| 21 | | | | | | 21 | | | | | |
| 22 | | | | | | 22 | | | | | |
| | | | | | | 23 | | | | | |
| | | | | | | 24 | | | | | |
| | | | | | | 25 | | | | | |
| | | | | | | 26 | | | | | |

***Table S2. Patient-antibody pairs*** *retrieved (in green), not performed (in gray) or excluded because of overly non-specific staining prohibiting proper segmentation (red).*

| | **I** | | **II** | **III** | **IV** | | **Total** |
|---|---|---|---|---|---|---|---|
| | Thickness (mm) | Longitudinal | mm | mm | mm | Circular | mm |
| **Control** | 1.5±0.9 | 10/17 | 2.6±1.5 | 18.3±6.5 | 2.3±0.7 | 11/16 | 23.5±7 |
| **Adenomyosis** | 1.2±0.8 | 10/19 | 2.3±1.2 | 19.5±6.0 | 2.2±1.7 | 10/16 | 23.7±6 |

***Table S3. Thickness and orientation of the 4 muscle layers of the myometrium.*** *The four layers are defined in Fig.1. Layer I was longitudinal (along the cervico-fundal axis) only in 50-60% of patients, and otherwise circular (wounding around the cervico-fundal axis). The orientation of layer II was always orthogonal to that of layer I. Layer III, the thickest layer, always had a twisted, variable orientation indicative of spirals, that cannot be clearly fathomed on a 2D section. Layer IV was circular in ~75% of patients, although the fibers often appeared elongated along the radial (serosa-to-endometrium) direction, making it hard to unambiguously define orientation of this layer. Variations in the orientation or layer thickness were not specific to the control or adenomyosis group. All thicknesses are indicated ± SD.*

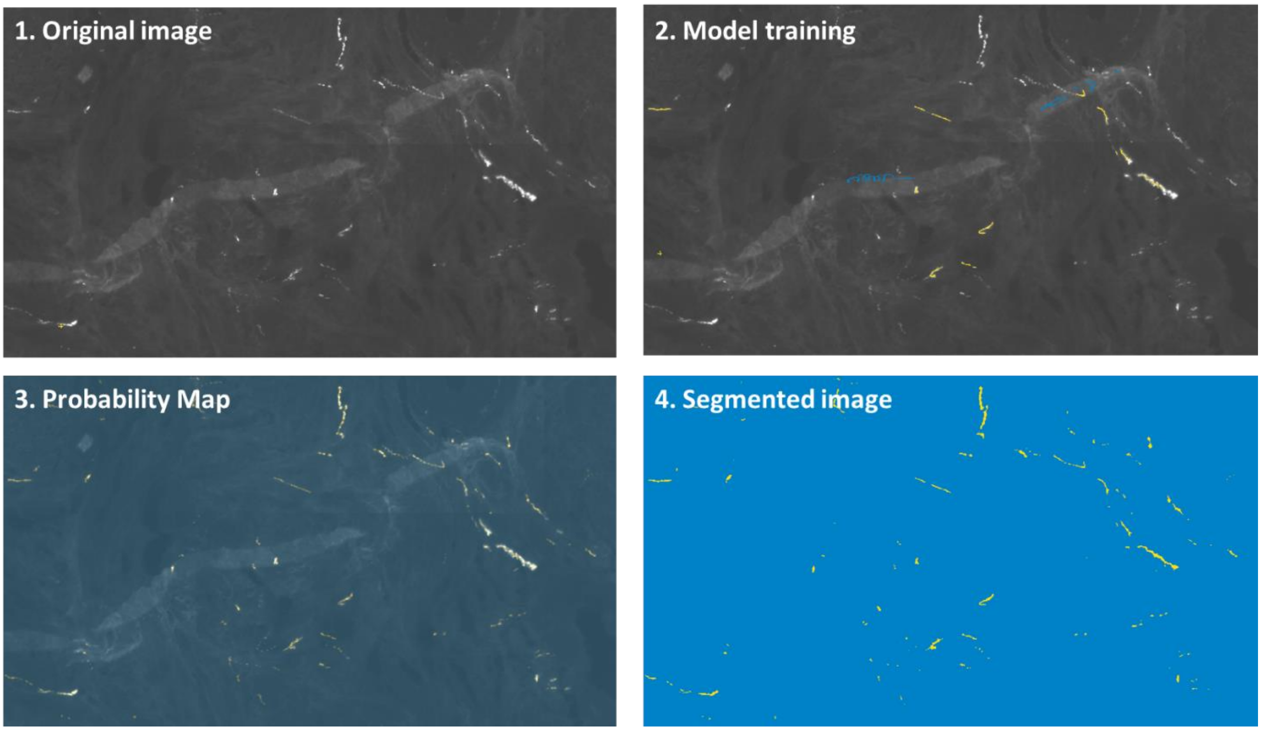


*Figure S1. Image analysis pipeline under Ilastik. The original image contains TH positive nerve fibers, but also some tissue folds with similar pixel intensity. The model is first trained by indicating what is a fiber (yellow) and what is not (blue). It then applies the model to the whole myometrium section, generating a probability map. A threshold on the minimum probability finally generates a segmented, binary image, that faithfully reproduces the area occupied by TH positive fibers.*

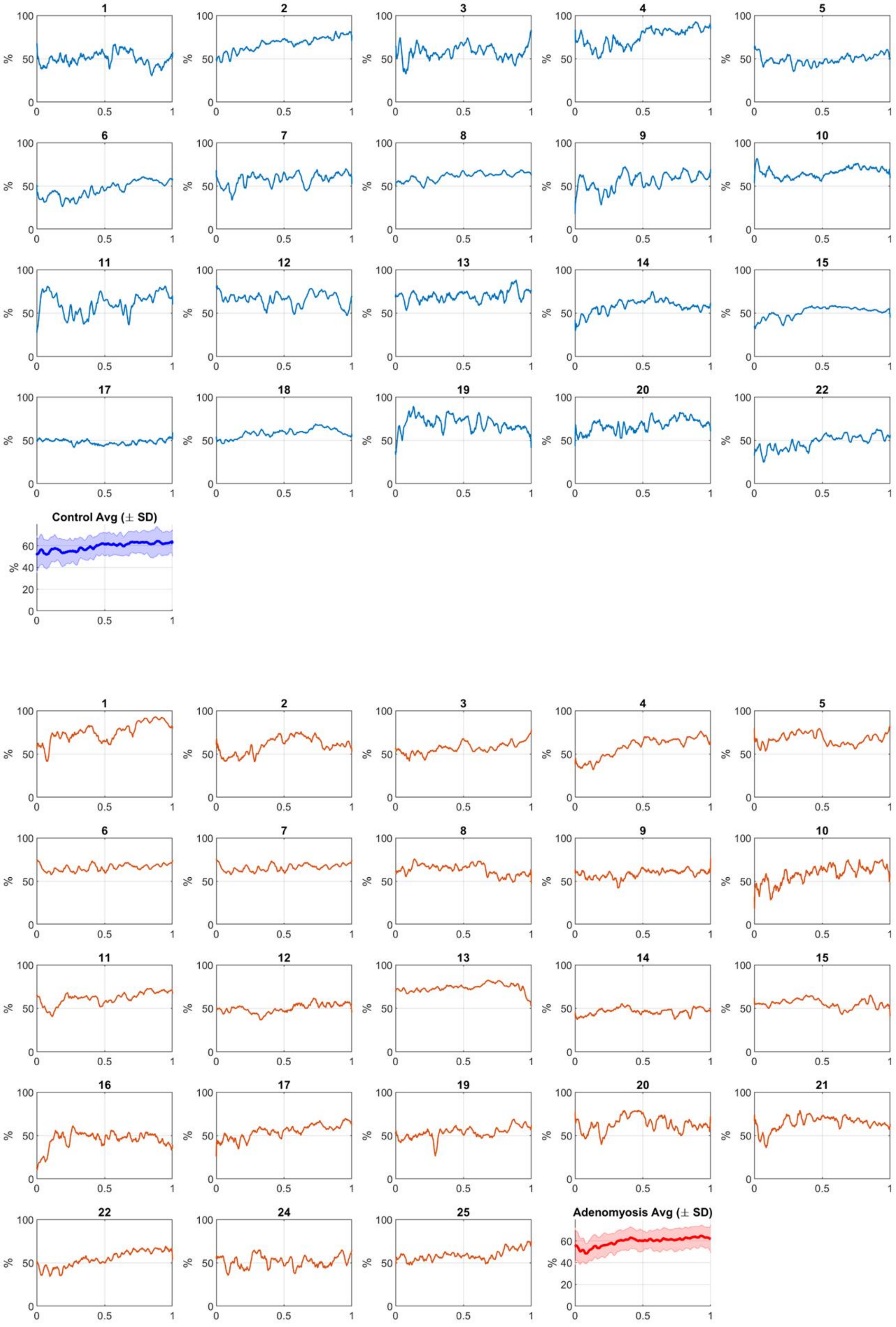


*Figure S2. α-SMA individual density profiles for controls (blue) and adenomyosis (red)*

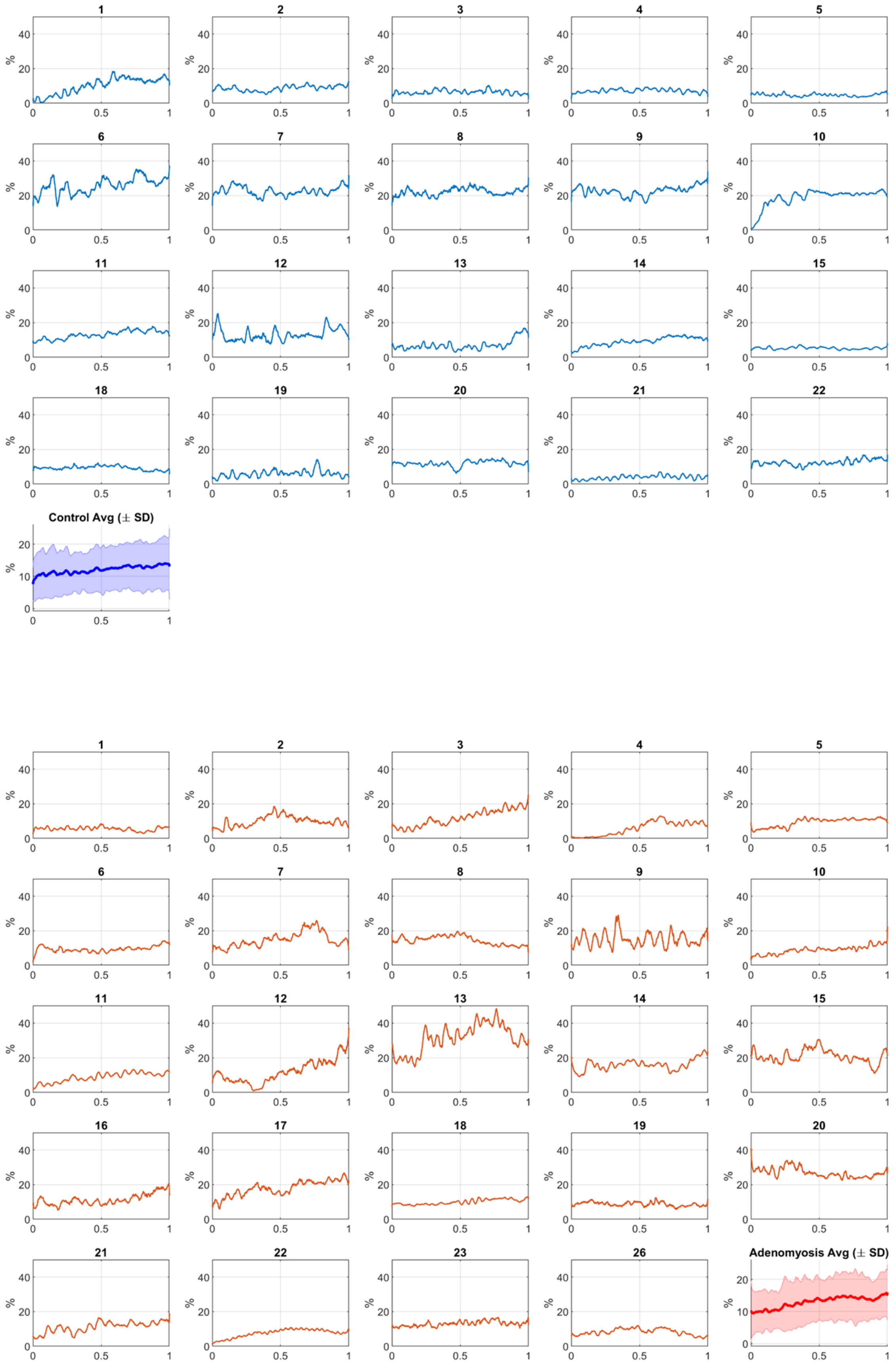


*Figure S3. PECAM individual density profiles for controls (blue) and adenomyosis (red)*

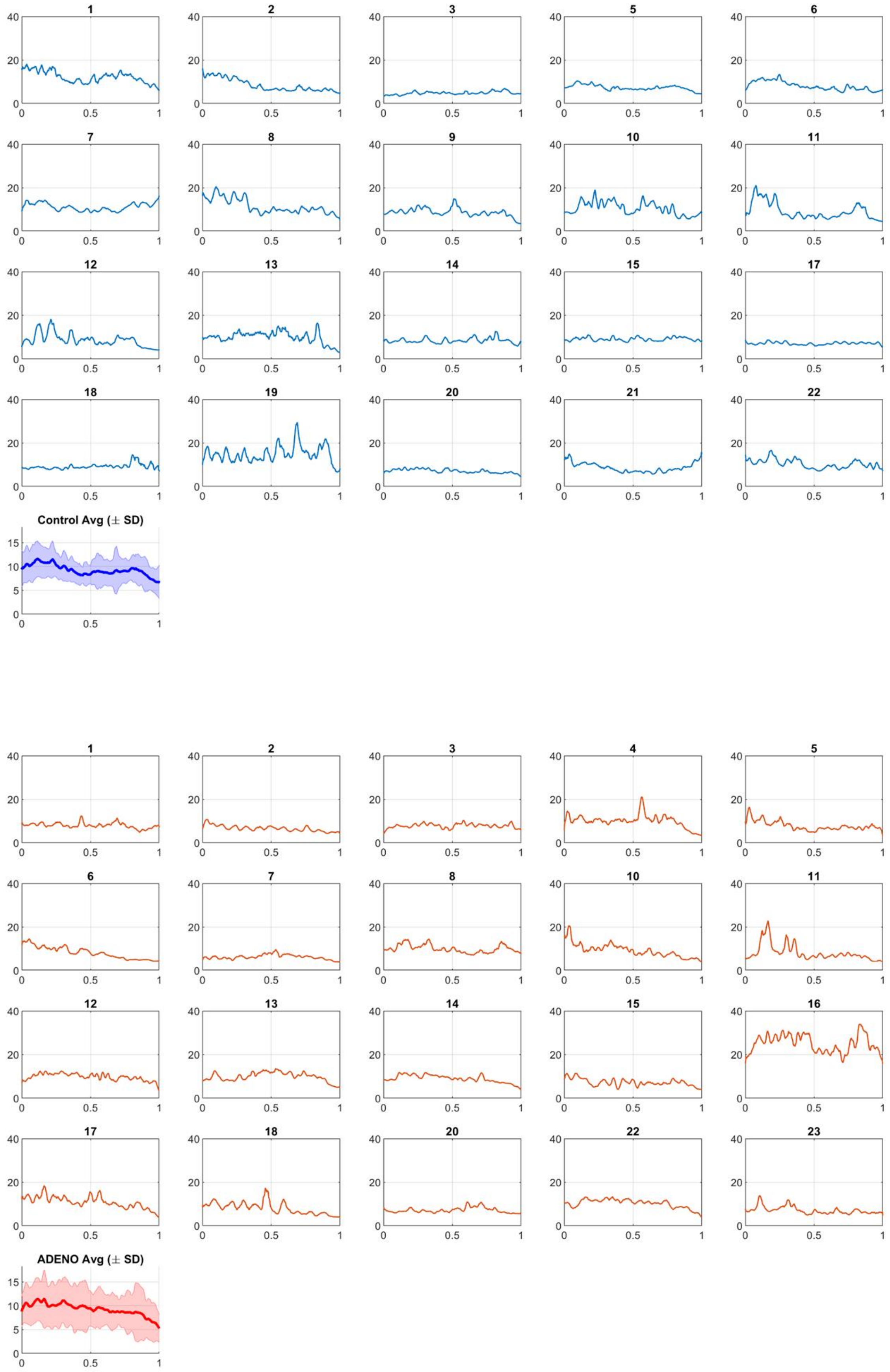


*Figure S4. SHG individual intensity profiles for controls (blue) and adenomyosis (red)*

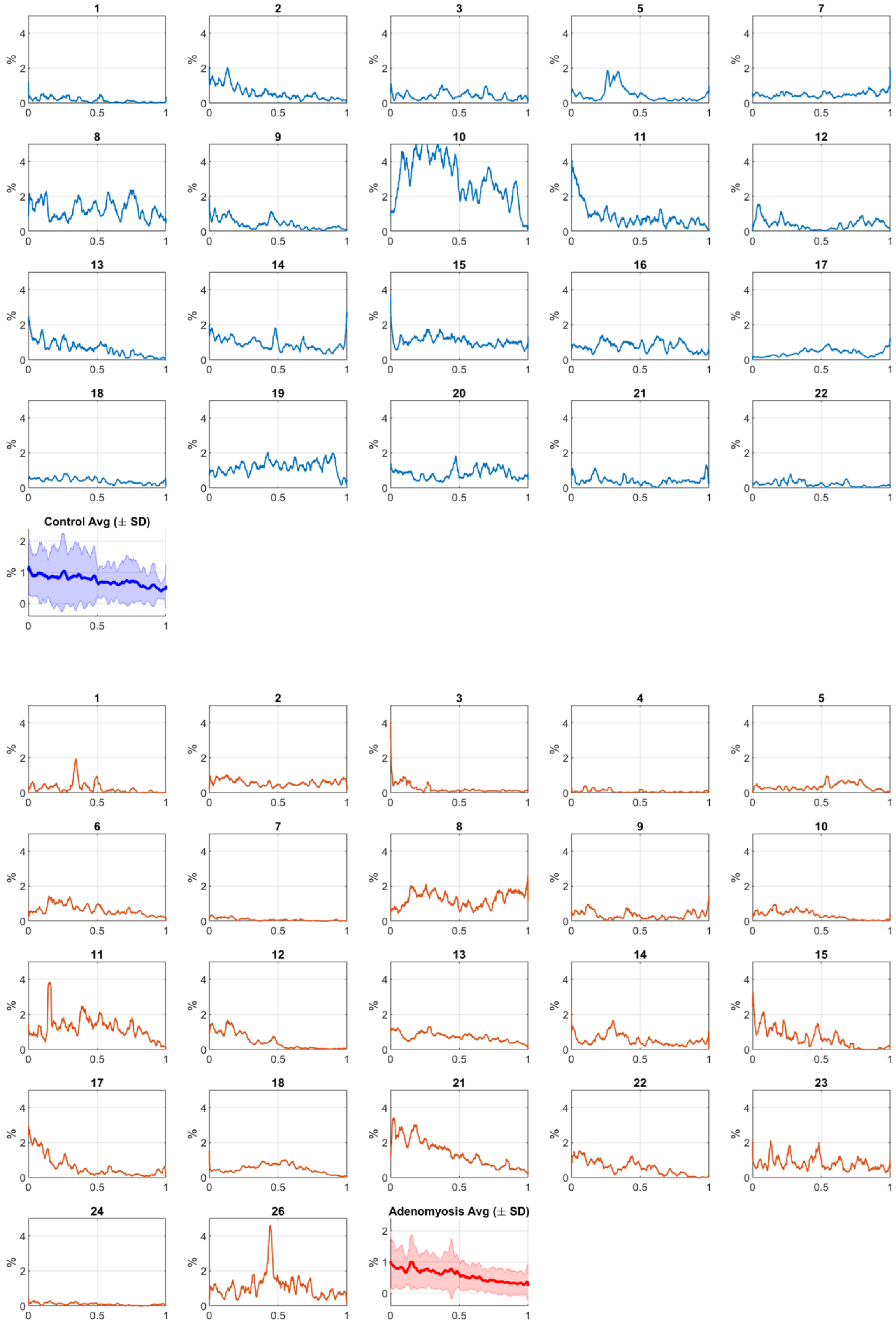


*Figure S5. PGP9.5 individual density profiles for controls (blue) and adenomyosis (red)*

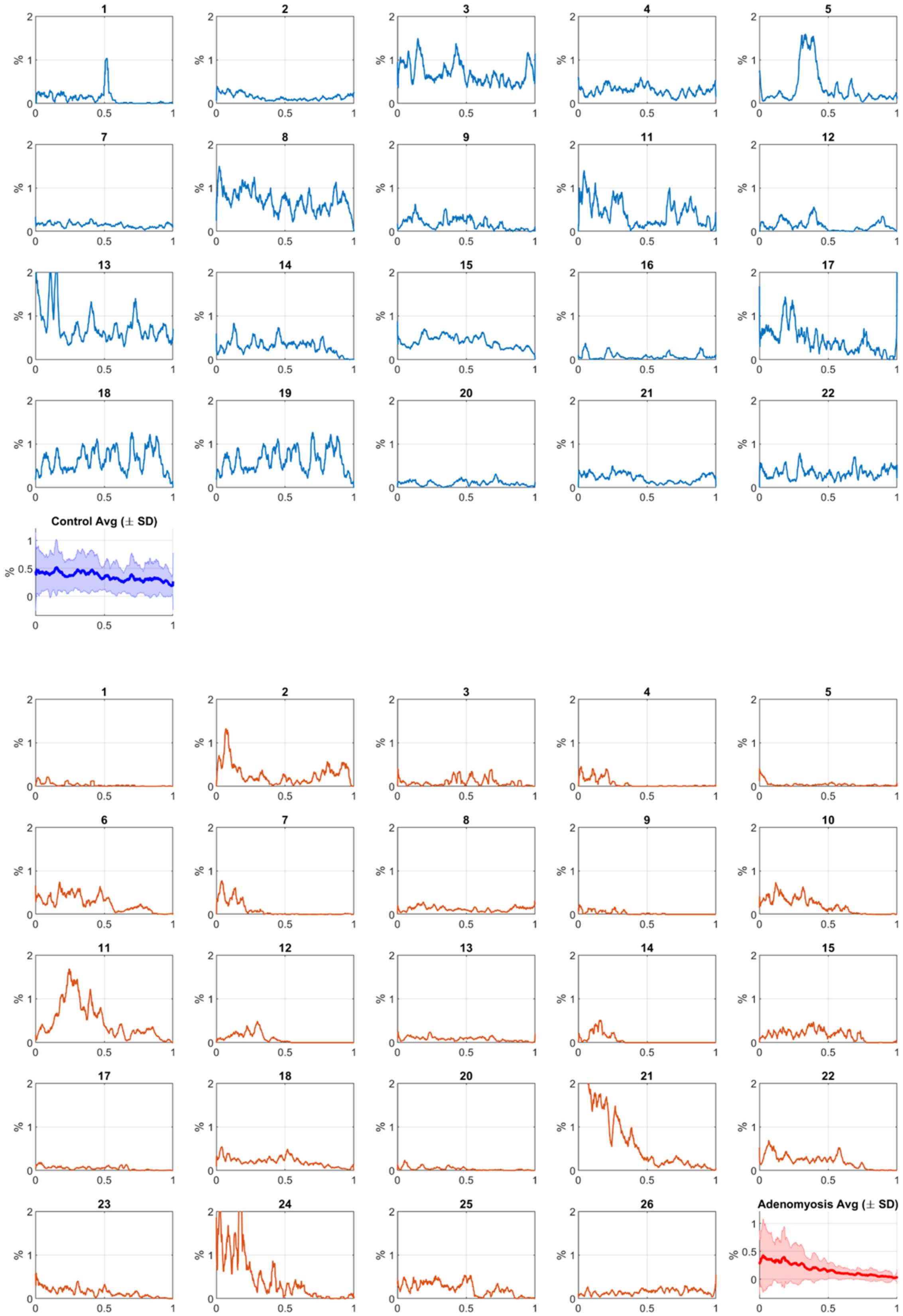


*Figure S6. TH individual density profiles for controls (blue) and adenomyosis (red)*

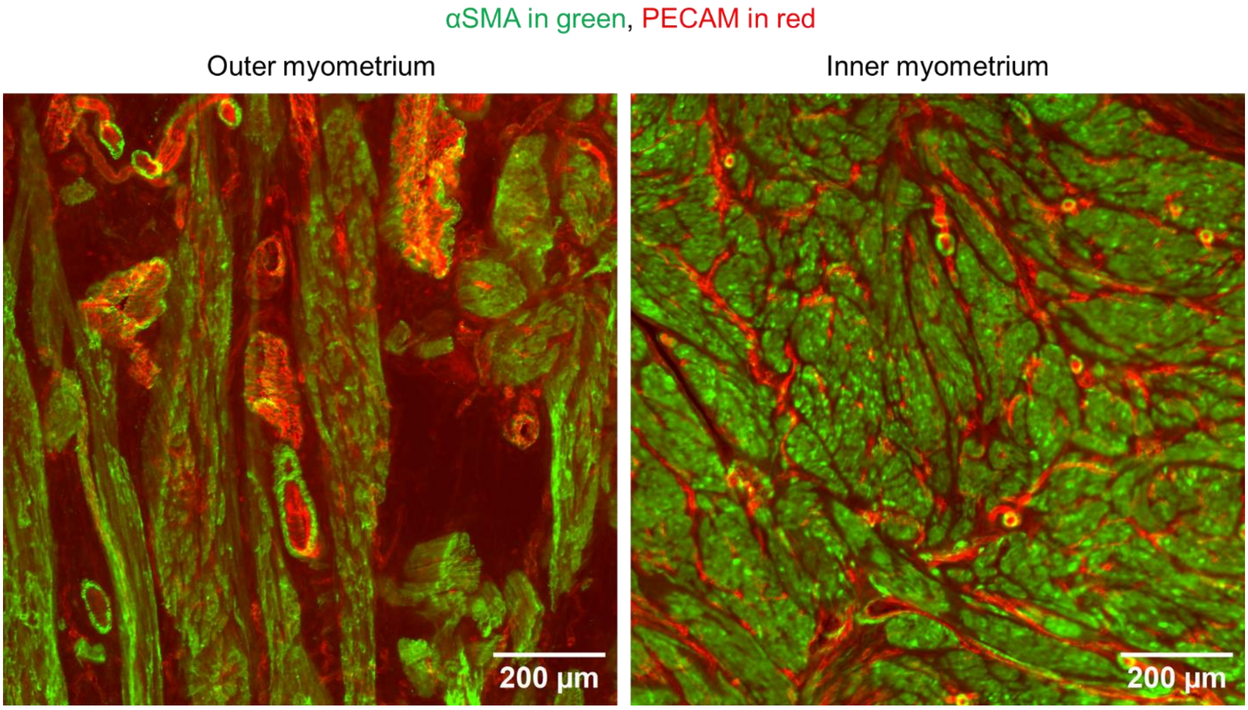


*Figure S7. Two-color immunofluorescence showing that most interstitial spaces between smooth muscle fibers are occupied by blood vessels in the inner, but not in the outer myometrium. Since PECAM only labels the endothelial lining, it does not directly yield the surface density occupied by blood vessels, which should additionally include blood vessel lumen and tunica. In the IM, most interstitial spaces between SM fibers were occupied by blood vessels (Fig.S5): the IM blood vessel density is likely ~30-40 %. In the OM, interstitial spaces are also occupied by other cell types or extracellular matrix.*

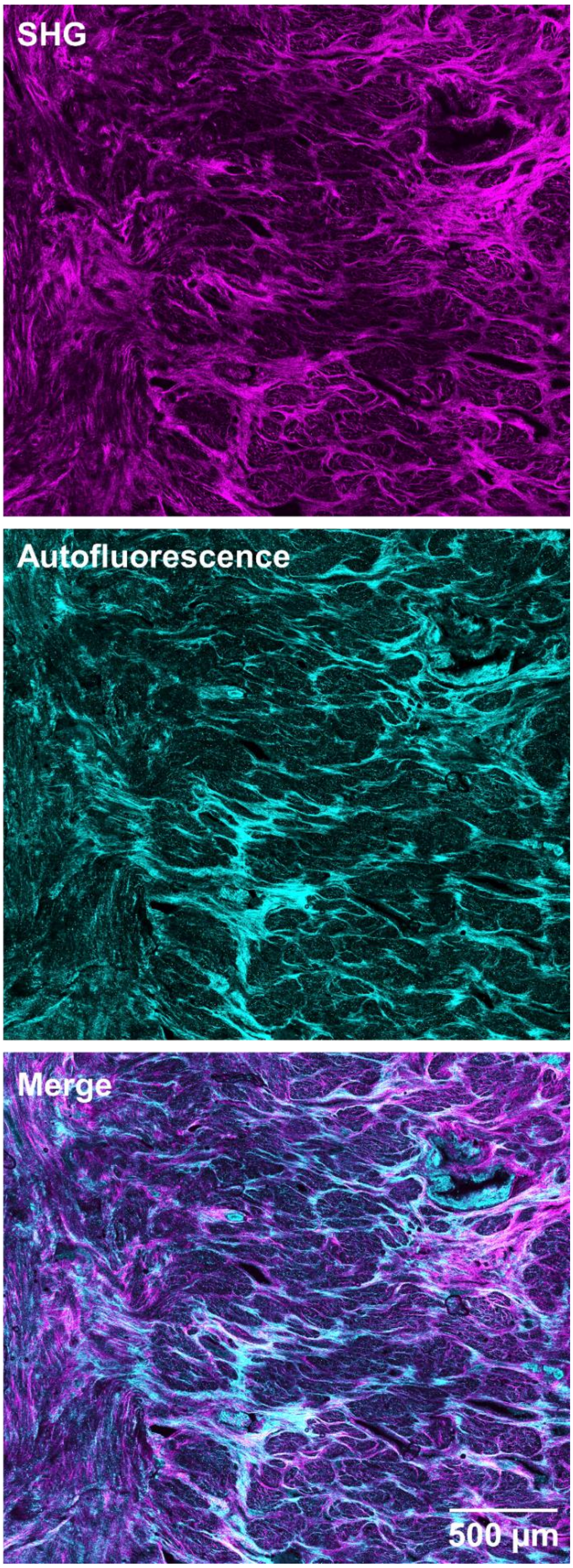


*Figure S8. The outer myometrium exhibits high GFP autofluorescence. It partially co-localizes with collagen fibers. Autofluoresence was likely due to elastin, which fluoresces in this wavelength domain (Bloksgaard et al., 2018), is often associated with collagen, and is known to be enriched in the OM (Kishi et al., 2017).*
